# Structural-functional multilayer brain network properties of the stimulation location predict outcome of repetitive transcranial magnetic stimulation for obsessive-compulsive disorder

**DOI:** 10.1101/2025.11.12.25340097

**Authors:** Sophie Fitzsimmons, Coen Coomans, Lucas Breedt, Neeltje Batelaan, Ysbrand van der Werf, Odile van den Heuvel, Linda Douw, Chris Vriend

## Abstract

**Background:** Repetitive transcranial magnetic stimulation (rTMS) is a promising treatment for obsessive-compulsive disorder (OCD), but response rates are variable. Pre-treatment characteristics of the stimulated region can influence rTMS treatment outcome.

**Objective/hypothesis:** We investigated the relationship between network features of the rTMS stimulation location and treatment outcome in a randomised trial of rTMS for OCD, using graph analysis of single-layer functional and structural brain networks, and of structural-functional multilayer networks.

**Methods:** We analysed data from 58 treatment-refractory adult OCD patients. Participants received either: high frequency (HF) rTMS to the left dorsolateral prefrontal cortex (DLPFC) (n=19); HF rTMS to the left pre-supplementary motor area (preSMA) (n=21); or control rTMS to the vertex (n=18). We used pre-treatment resting state functional MRI (rs-fMRI) and diffusion MRI (dMRI) scans to construct single-layer and structural-functional multilayer networks for each participant. We computed various centrality measures of the stimulated location and connected subnetwork, and examined their relationship with treatment outcome.

**Results:** We found no associations between functional or structural single-layer network characteristics and treatment outcome. However, higher average multilayer betweenness centrality of the stimulated subnetwork in the DLPFC rTMS group (but not in the preSMA or vertex groups) was associated with greater symptom reduction (p=0.013).

**Conclusions:** Participants with greater integration of the stimulated subnetwork with the rest of the structural-functional network showed greater improvement following DLPFC rTMS. Our results give a preliminary indication that the outcome of rTMS treatment for OCD is dependent on the interplay between structural and functional networks.

## Introduction

Repetitive transcranial magnetic stimulation (rTMS) is an increasingly prevalent non-invasive brain stimulation treatment technique in psychiatry, primarily used for depression, but also showing promise for several other disorders, including obsessive-compulsive disorder (OCD)[1–3]. RTMS works by delivering a train of magnetic pulses to a specific brain region, inducing neuroplasticity: changes in brain structure and function at both the stimulation location and distant connected regions that are related to clinical improvements[4,5]. Response to rTMS is, however, variable, partly due to inter-individual variation in the neurobiological properties of the targeted brain area and (sub)network. There is substantial evidence from studies of rTMS treatment for depression that the functional connectivity (FC) (or correlation between activity fluctuations) of the stimulation location with other connected brain regions has an influence on rTMS response: for example, FC of the dorsolateral prefrontal cortex (DLPFC) with the subgenual anterior cingulate cortex[6–9] is associated with rTMS treatment outcome. There is also evidence to suggest that structural, white-matter connectivity is a mediator of rTMS effect: diffusion MRI (dMRI) measures such as fractional anisotropy or streamline count close to the stimulated area have been found to be related to rTMS outcome in depression[10–12].

Connectivity is only one brain feature related to rTMS response: the spatial organisation or topology of the brain network may also be relevant. The topology of the brain network can be described using graph/network analysis, where nodes of the network are brain areas, defined by an atlas, and their connections are defined by the correlation in activity between nodes (for functional MRI (fMRI)-derived connectomes) or by the number of streamlines (for dMRI-derived structural connectomes)[13]. Graph metrics describe the organisation of the network or the role of a specific node in the network: for example, centrality measures indicate the extent to which a node can influence or be influenced by other nodes by describing various aspects of its connectivity[14]. Studies of single-session rTMS in both controls[15] and OCD patients[16] using centrality graph measures extracted from fMRI networks have shown that greater baseline local connectivity (i.e. connectivity to neighbouring nodes) and less integration (i.e. connectivity throughout the entire network) of the stimulation site and stimulated network is associated with greater rTMS effects. The functional brain network is, however, dependent on and constrained by the structural network[17–19]: examining the combined contributions of functional and structural connectivity may have greater explanatory value than either type of connectivity alone[20,21]. One method of examining the combination of brain structure and function is by constructing multilayer networks[20,22,23], which consist of multiple linked networks with the same nodes but different edge definitions (for example, a two-layer network with links defined by rsfMRI and by dMRI). It is thought that multilayer brain networks capture the interplay between different network layers that analysing modalities separately may miss[22,24], and allow the integration of multiple sources of information about network organisation. A recent study by Breedt et al[20] showed that while centrality of single layer rsfMRI-, dMRI- or resting-state MEG-derived networks did not predict executive function in controls, centrality of the multilayer network did. Other studies have highlighted the clinical relevance of multilayer networks, showing that multilayer network features can discriminate between healthy controls and Alzheimer’s disease[25] and schizophrenia[26], and track longitudinal changes in executive function in glioma patients[27].

In this study, we used neuroimaging data from a recent clinical trial of combined rTMS and exposure and response prevention psychotherapy (ERP) for OCD[28] to examine the association between treatment outcome and rsfMRI- and dMRI-derived single- and multilayer network features of the stimulated network and stimulation location.

We investigated:

1. whether baseline graph features of functional and structural single-layer networks were associated with treatment outcome.
2. whether baseline graph features of a structural-functional multilayer network were associated with treatment outcome.

For single-layer functional networks, we hypothesised that higher local connectivity and lower global connectivity would predict greater response to rTMS. We expected to see the same direction of effect for structural as for functional graph features, because structural connectivity is generally correlated with FC[19] (though this can vary across the cortex[29]), and studies examining other dMRI connectivity features[10–12] also suggest that greater local connectivity may predict greater rTMS effects. There is no literature on the relationship between rTMS effects and multilayer network organisation. Given that structural-functional multilayer networks summarise the combined contribution of structural and functional graph features, we hypothesised the same directions of effect as for single-layer graph measures.

## Material and Methods

### Participants

This study uses data from the TIPICCO trial (TMS-induced plasticity improving cognitive control in OCD, registered at NCT03667807). Full methods are reported elsewhere[28]. Sixty-four adults with OCD started the treatment. Eligibility criteria were: meeting DSM-5 criteria for OCD as determined by the Structured Clinical Interview for DSM-5[30], moderate/severe OCD symptoms (Yale-Brown Obsessive-Compulsive Scale[31] (YBOCS) score of ≥16), age 18-65 years, previous treatment with ≥8 sessions of ERP or cognitive-behavioural therapy for OCD, and ≥12 weeks of previous serotonin reuptake inhibitor (SRI) treatment for OCD or medication naïve due to a strong preference for non-pharmacological treatment. Exclusion criteria were: Tourette’s syndrome, schizophrenia, bipolar disorder, active suicidal ideation, MRI/rTMS exclusion criteria, and previous experience with rTMS. Participants were allowed to continue current SRI medication at the same dose. The study was approved by the ethics committee of the VU Medical Center (VUmc), Amsterdam, The Netherlands, and was conducted as a collaboration between the VUmc and the mental health care institute GGZ inGeest, Amsterdam. All participants provided written informed consent in accordance with the declaration of Helsinki.

### Study design

This was a three-arm parallel randomised controlled trial, conducted between May 2019 and October 2022. Participants were randomised to one of three types of rTMS (10Hz, targeting either L DLPFC or L pre-supplementary motor area [preSMA] at 110% motor threshold, or vertex at 60% motor threshold as a control condition), all immediately followed by ERP. This was delivered twice a week, for 8 weeks (20 treatment sessions in total, of which 16 were rTMS-ERP sessions and 4 ERP only). Individualised coordinates for neuronavigated targeting of rTMS for the DLPFC and preSMA groups were determined using task-based fMRI acquired at baseline (T0) (during the Tower of London task for the DLPFC, and the Stop-signal task for the preSMA) (see Supplement and [28] for full details of rTMS and ERP treatments, and rTMS target determination). OCD symptom severity was assessed using the YBOCS at T0, after rTMS-ERP session 8 (T1), after rTMS-ERP session 16 (T2), and 12 weeks following the end of treatment (T3); only measurements from T0 and T2 are used in this study. Participants also underwent multimodal MRI scans at T0 and T2 (see *Image acquisition* below); only dMRI and rsfMRI scans at T0 are analysed in this study.

### Image acquisition

Participants were scanned on a GE Signa HDxT 3T MRI scanner using a 32-channel head coil (General Electric, Milwaukee, U.S). Anatomical MRI was acquired using a 3D T1-weighted structural magnetization-prepared rapid acquisition gradient-echo (MPRAGE; TR = 6.9 ms; TI = 900 ms; TE = 3.0 ms; 256x256 matrix; 1 mm3 isotropic resolution; 168 sections) and was used for coregistration and normalisation of fMRI and dMRI scans. All rsfMRI scans were acquired using a gradient echo-planar imaging sequence (TR = 2.2 s; TE = 26 ms; 64x64 matrix; field of view 21.1cm; flip angle = 80°) with 42 ascending slices per volume (3.3 x 3.3 mm in-plane resolution; slice thickness = 3.0 mm; interslice gap = 0.3 mm). Participants were scanned in a darkened room, and were instructed to remain awake with their eyes closed during the scan. DMRI multi-shell images consisted of 73 diffusion-weighted directions (25 b1000, 24 b2000, and 24 b3000 s/mm^2^) and seven interleaved non-diffusion-weighted volumes (b0 s/mm^2^). Additional fieldmaps (for both the rsfMRI and dMRI) were acquired with opposite phase-encoding directions for distortion correction.

### rsfMRI processing

RsfMRI data were pre-processed using fMRIprep v21.0.1 (https://fmriprep.org) (see Supplement for boilerplate). In fMRIprep, reversed phase-encoding scans were used to account for susceptibility-induced distortions, volume alignment was carried out, and motion parameters were extracted. Additional steps included motion correction (performed using Automatic Removal of Motion Artifacts and Independent Component Analysis (ICA-AROMA)[32] and regressing out sources of physiological noise from grey and white matter), 0.009-0.08Hz bandpass filtering, and 6mm smoothing using FSL SUSAN[33]. Parcellation of each participant’s T1-weighted image was carried out using the Schaefer 300 cortical atlas[34] plus 14 subcortical regions individually segmented using FreeSurfer (v7.1.1)[35]. This was followed by extraction of average BOLD-signals from each brain area. The z-transformed, absolutised Pearson correlations between each node’s timeseries formed the weighted edges of the functional network.

### dMRI processing

Diffusion images were denoised using the *dwidenoise* tool in MRtrix3[36], and corrected for eddy currents, susceptibility-induced distortions, and within- and between-volume motion using eddy from the FMRIB Software Library (FSL; v6.0.5). We used eddyqc[37] to extract image quality measures. Outliers were visually inspected to identify volumes with residual (motion-related) artefacts and deleted if necessary. Structural connectomes were constructed using probabilistic anatomically-constrained tractography (ACT)[38] in MRtrix3[36]. Tissue response function estimation was carried out using the multishell multitissue five-tissue-type algorithm (msmt_5tt), followed by multishell multitissue constrained spherical deconvolution (MSMT-CSD)[39]. Tractograms were constructed by random seeding of 50 million fibres at the grey matter/white matter boundary, followed by spherical-deconvolution informed filtering of tractograms (SIFT2 from MRtrix3)[40]. Finally, the above parcellation of each participant’s T1-weighted image was used to convert the tractogram to a structural network, where weighted links represented streamline counts between all voxels within two brain regions.

### Parcellation and definition of whole-brain atlas, stimulated node and subnetwork

Due to signal loss in the rsfMRI data, we were not able to retain all atlas regions; for our analyses we used the subset of atlas nodes common to both the structural and functional data of all participants. Our final atlas consisted of 307 nodes (we excluded 7 nodes in bilateral orbital frontal cortex and bilateral temporal pole: see Supplement for specific regions).

Stimulated nodes were defined as the atlas region that overlapped most with a 5mm radius sphere constructed around the stimulated rTMS coordinates in MNI space. For the vertex group, the DLPFC and preSMA coordinates were defined in the same way as in the DLPFC and preSMA groups to act as control comparison coordinates (further referred to as vertex-DLPFC and vertex-preSMA, respectively), but these participants were not stimulated at these locations. The stimulated subnetwork was defined as the Yeo 7-network subdivision[41] containing the stimulated node.

### Graph analyses and network measure extraction

#### Single-layer networks

Weighted networks were constructed for both structural and functional data. The following graph measures were calculated using the Brain Connectivity Toolbox (BCT)[42](https://sites.google.com/site/bctnet/):

- <u>Nodal measures:</u> node strength (NS), betweenness centrality (BC), participation coefficient (PC) and eigenvector centrality (EC) (see Box 1) of stimulated and hypothetical stimulated nodes in rsfMRI and dMRI networks

- <u>Subnetwork measures</u>: NS, BC, PC and EC of all nodes in each stimulated and hypothetically stimulated Yeo subnetwork and averaged across the subnetwork.

##### Box 1

###### Graph measure definitions

- <u>Node strength</u> - the total strength of a nodés connections, or how strongly connected a node is to its neighbours. Can be considered a measure of local connectivity[42].

- <u>Degree centrality</u> - the equivalent of node strength for binarised networks: the number of connections a node has.

- <u>Betweenness centrality</u> - measures how many high strength paths in the network pass through a node, giving a measure of global connectivity (connectivity throughout the entire network)[42].

- <u>Participation coefficient</u> - a high participation coefficient indicates a relatively high number of connections outside compared to within a node’s subnetwork[43].

- <u>Eigenvector centrality</u> - denotes the influence of a node in a network - high EC nodes have connections with nodes that also connect with highly central nodes[44].

#### Multilayer networks

Binary multiplex networks were constructed, using the same in-house scripts as Breedt et al 2023[20] (https://github.com/multinetlab-amsterdam/projects/tree/master/mumo_paper_2021), with two layers consisting of 1) a functional, rsfMRI-derived minimum spanning tree (MST) network (MSTs are a type of binarised network, formed from the strongest links of the weighted version of the network, with links of weight=1 that connect all the nodes in the network without forming loops) and 2) a structural, dMRI-derived MST network. Interlayer links were present only between the same nodes across layers (with weight=1).

The following graph measures were calculated:

- <u>Nodal measures</u>: degree centrality (DC), BC, and EC of stimulated and hypothetical stimulated nodes in multilayer networks were extracted using in-house scripts (https://github.com/multinetlab-amsterdam/data_analysis/tree/Multilayer/Multilayer).

- <u>Subnetwork measures</u>: DC, BC, and EC of all nodes in each stimulated and hypothetically stimulated Yeo subnetwork were extracted and averaged across the subnetwork.

### Statistical analyses

Analyses were pre-registered at https://osf.io/s9krj. All statistical analyses were performed in R (v4.2.1, Vienna, Austria).

Primary outcome: We performed linear regression analyses to statistically test the association between baseline graph measures and treatment response, as defined by pre-post treatment change in YBOCS score (ΔYBOCS). The node and subnetwork graph measures NS, BC, PC, and EC in structural and functional single-layer networks, and DC, BC, and EC for multilayer networks, were included in separate models as the dependent variable. The independent variable was defined as ΔYBOCS, with covariates age, sex and baseline YBOCS. Analyses were performed for each group separately (DLPFC, preSMA, vertex-DLPFC, vertex-preSMA). To correct for multiple tests, and since graph measures are highly correlated, we used a Sidak/D-AP-corrected alpha of 0.019 for the single layer networks and 0.025 for the multilayer networks, calculated via the SISA website (https://www.quantitativeskills.com/sisa/calculations/bonfer.htm) based on the average correlation between the graph measures (r = 0.301 for single-layer networks, r=0.361 for multilayer networks). We also performed these analyses in all groups combined in order to examine general associations with treatment outcome.

Exploratory outcomes: Because the preSMA and DLPFC coordinates were individually determined, different Yeo resting state subnetworks[41] were stimulated in each participant. We therefore examined whether symptom improvement following rTMS was dependent on the subnetwork that the stimulated node happened to occur in. Participants in the preSMA and DLPFC groups were combined and then regrouped according to which Yeo subnetwork was stimulated. ΔYBOCS was compared between stimulated Yeo subnetworks using a t-test and responder rates using a chi-squared test. Subnetworks containing the hypothetically stimulated nodes in the vertex group were compared separately as a control. Demographic characteristics were compared between groups using one-way ANOVAs, chi-squared tests, or Kruskal-Wallace tests as appropriate. ΔYBOCS over time was compared between groups using linear mixed models (see also [28]).

## Results

### Demographic information

Of the 64 participants that started treatment, three participants dropped out during treatment and lacked clinical data at T2, two did not have rsfMRI/dMRI scans available, and one was excluded due to lack of fMRI signal at the stimulation location. Fifty-eight had usable rsfMRI/dMRI data at T0 and clinical measurements at both T0 and T2 and are included in the present study (19 in the DLPFC group, 21 in the preSMA group, 18 in the vertex group). Clinical and demographic details are presented in Table 1. Groups were generally well-matched across demographic and baseline clinical characteristics, though the preSMA group was significantly younger than the other groups (p=0.004). The vertex group had a lower Beck Depression Index (BDI) score than the other groups (p=0.003), and lower rates of comorbid depression (p=0.029).

**Table 1:**
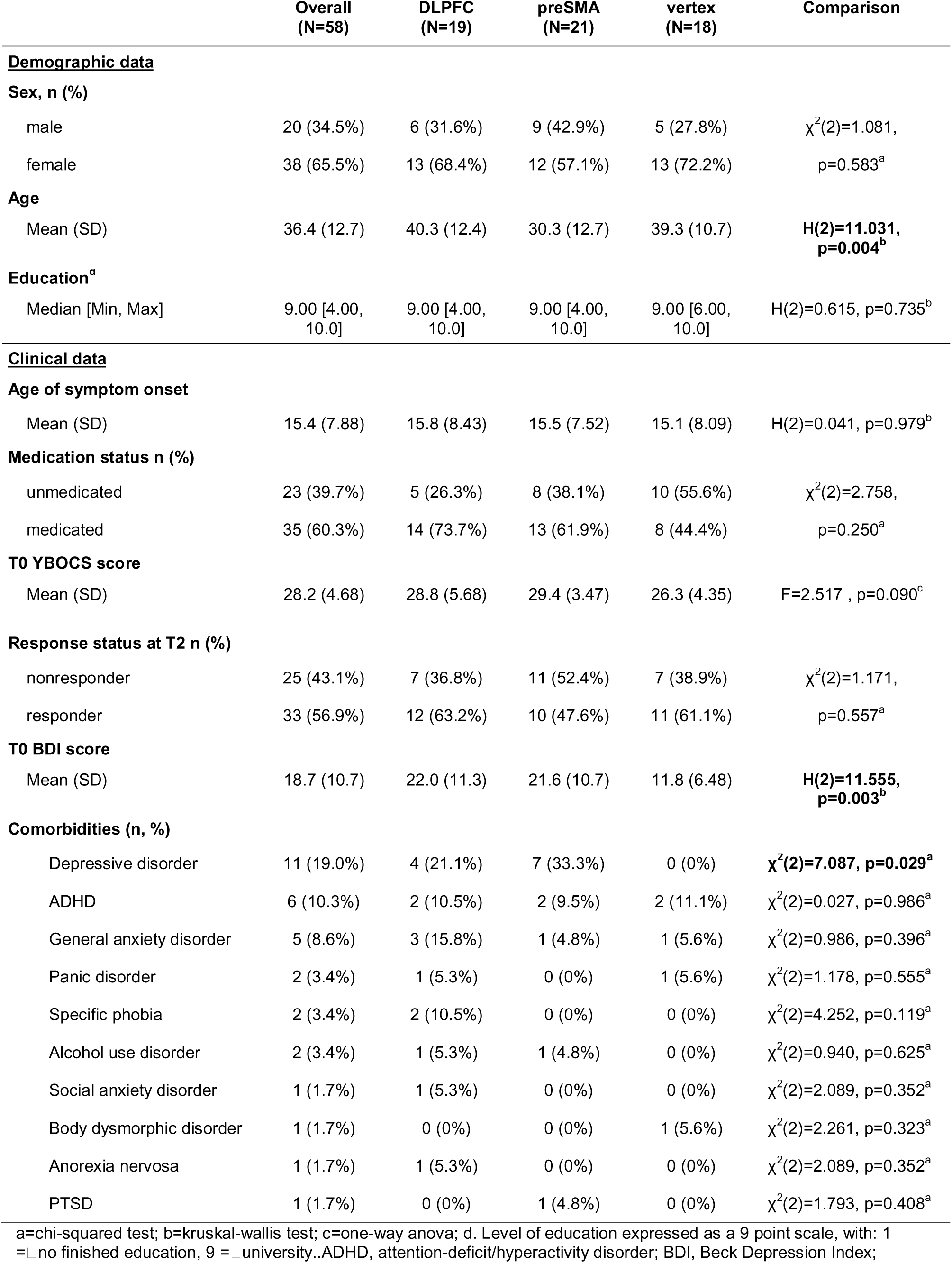
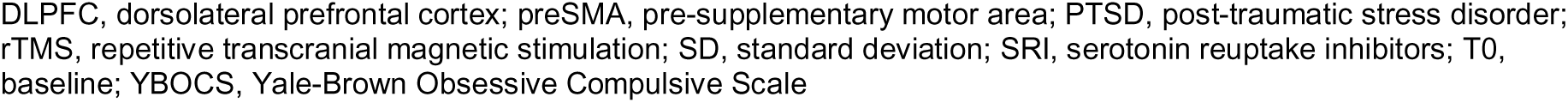
demographic and clinical information.

|  | Overall<br>(N=58) | DLPFC<br>(N=19) | preSMA<br>(N=21) | vertex<br>(N=18) | Comparison |
| --- | --- | --- | --- | --- | --- |
| **Demographic data** |  |  |  |  |  |
| **Sex, n (%)** |  |  |  |  |  |
| male | 20 (34.5%) | 6 (31.6%) | 9 (42.9%) | 5 (27.8%) | $\chi^2(2)=1.081$,<br>p=0.583<sup>a</sup> |
| female | 38 (65.5%) | 13 (68.4%) | 12 (57.1%) | 13 (72.2%) |  |
| **Age** |  |  |  |  |  |
| Mean (SD) | 36.4 (12.7) | 40.3 (12.4) | 30.3 (12.7) | 39.3 (10.7) | **H(2)=11.031**,<br>**p=0.004<sup>b</sup>** |
| **Education<sup>d</sup>** |  |  |  |  |  |
| Median [Min, Max] | 9.00 [4.00, 10.0] | 9.00 [4.00, 10.0] | 9.00 [4.00, 10.0] | 9.00 [6.00, 10.0] | H(2)=0.615, p=0.735<sup>b</sup> |
| **Clinical data** |  |  |  |  |  |
| **Age of symptom onset** |  |  |  |  |  |
| Mean (SD) | 15.4 (7.88) | 15.8 (8.43) | 15.5 (7.52) | 15.1 (8.09) | H(2)=0.041, p=0.979<sup>b</sup> |
| **Medication status n (%)** |  |  |  |  |  |
| unmedicated | 23 (39.7%) | 5 (26.3%) | 8 (38.1%) | 10 (55.6%) | $\chi^2(2)=2.758$,<br>p=0.250<sup>a</sup> |
| medicated | 35 (60.3%) | 14 (73.7%) | 13 (61.9%) | 8 (44.4%) |  |
| **T0 YBOCS score** |  |  |  |  |  |
| Mean (SD) | 28.2 (4.68) | 28.8 (5.68) | 29.4 (3.47) | 26.3 (4.35) | F=2.517 , p=0.090<sup>c</sup> |
| **Response status at T2 n (%)** |  |  |  |  |  |
| nonresponder | 25 (43.1%) | 7 (36.8%) | 11 (52.4%) | 7 (38.9%) | $\chi^2(2)=1.171$,<br>p=0.557<sup>a</sup> |
| responder | 33 (56.9%) | 12 (63.2%) | 10 (47.6%) | 11 (61.1%) |  |
| **T0 BDI score** |  |  |  |  |  |
| Mean (SD) | 18.7 (10.7) | 22.0 (11.3) | 21.6 (10.7) | 11.8 (6.48) | **H(2)=11.555**,<br>**p=0.003<sup>b</sup>** |
| **Comorbidities (n, %)** |  |  |  |  |  |
| Depressive disorder | 11 (19.0%) | 4 (21.1%) | 7 (33.3%) | 0 (0%) | **$\chi^2(2)=7.087$, p=0.029<sup>a</sup>** |
| ADHD | 6 (10.3%) | 2 (10.5%) | 2 (9.5%) | 2 (11.1%) | $\chi^2(2)=0.027$, p=0.986<sup>a</sup> |
| General anxiety disorder | 5 (8.6%) | 3 (15.8%) | 1 (4.8%) | 1 (5.6%) | $\chi^2(2)=0.986$, p=0.396<sup>a</sup> |
| Panic disorder | 2 (3.4%) | 1 (5.3%) | 0 (0%) | 1 (5.6%) | $\chi^2(2)=1.178$, p=0.555<sup>a</sup> |
| Specific phobia | 2 (3.4%) | 2 (10.5%) | 0 (0%) | 0 (0%) | $\chi^2(2)=4.252$, p=0.119<sup>a</sup> |
| Alcohol use disorder | 2 (3.4%) | 1 (5.3%) | 1 (4.8%) | 0 (0%) | $\chi^2(2)=0.940$, p=0.625<sup>a</sup> |
| Social anxiety disorder | 1 (1.7%) | 1 (5.3%) | 0 (0%) | 0 (0%) | $\chi^2(2)=2.089$, p=0.352<sup>a</sup> |
| Body dysmorphic disorder | 1 (1.7%) | 0 (0%) | 0 (0%) | 1 (5.6%) | $\chi^2(2)=2.261$, p=0.323<sup>a</sup> |
| Anorexia nervosa | 1 (1.7%) | 1 (5.3%) | 0 (0%) | 0 (0%) | $\chi^2(2)=2.089$, p=0.352<sup>a</sup> |
| PTSD | 1 (1.7%) | 0 (0%) | 1 (4.8%) | 0 (0%) | $\chi^2(2)=1.793$, p=0.408<sup>a</sup> |

a=chi-squared test; b=kruskal-wallis test; c=one-way anova; d. Level of education expressed as a 9 point scale, with: 1 = no finished education, 9 = university..ADHD, attention-deficit/hyperactivity disorder; BDI, Beck Depression Index;
DLPFC, dorsolateral prefrontal cortex; preSMA, pre-supplementary motor area; PTSD, post-traumatic stress disorder; rTMS, repetitive transcranial magnetic stimulation; SD, standard deviation; SRI, serotonin reuptake inhibitors; T0, baseline; YBOCS, Yale-Brown Obsessive Compulsive Scale

### Clinical outcomes

Full clinical outcomes are reported elsewhere[28]. Symptom severity decreased significantly at T2 compared to T0 (T0-T2 mean difference = -10.836, p<0.001, 95% CI [-12.504, -9.168])). There was no statistically significant group by time interaction, both before and after adjusting for age, sex, and baseline BDI, indicating no group differences in symptom reduction.

### Distribution of stimulation locations

See Figure 1 for a visualisation of the stimulation locations and the stimulated Yeo resting state networks. The stimulation location for the majority of the DLPFC group was in the frontoparietal network (FPN, 14/19) while for the preSMA group this was the default mode network (DMN, 16/21). There was no significant difference in symptom improvement (ΔYBOCS) (t(37)=1.01, p=0.318) or number of responders (χ^2^(1)=0.710, p=0.400) between those stimulated in the FPN and those stimulated in the DMN.

**Figure 1:**
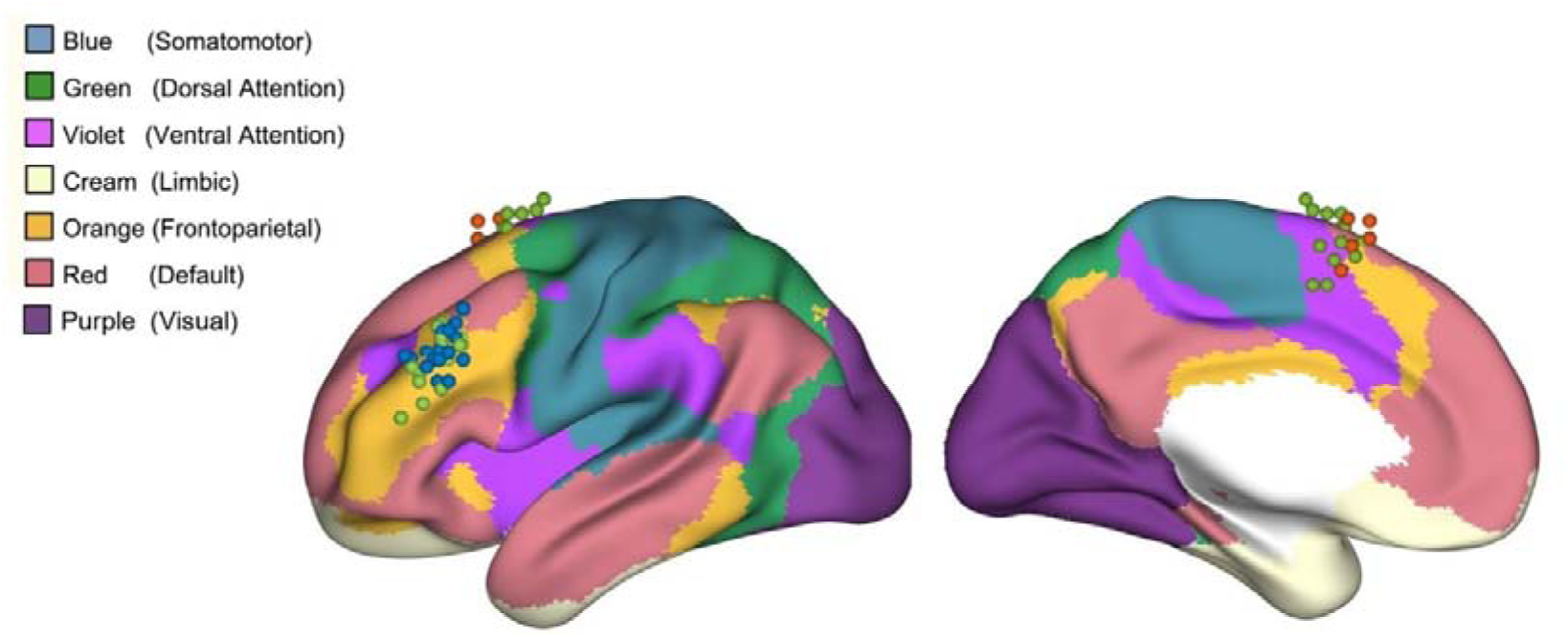
lateral (left) and medial (right) view of rTMS stimulation locations and their localisation within Yeo resting state networks. Dorsolateral prefrontal cortex group stimulation locations (n=19) in blue; pre-supplementary motor area group (n=23) in red; hypothetical preSMA and DLPFC stimulation locations for vertex group (n=18) in light green.

### Prediction of treatment outcome using single-layer network properties

Following correction for multiple comparisons, no single-layer structural or functional graph features were associated with treatment outcome in either active rTMS or control groups (See supplementary tables S1 and S2 for multiple regression results). We also found no significant associations between graph features and treatment outcome in all participants combined for either type of single-layer network.

### Prediction of treatment outcome using multilayer network properties

Greater multilayer BC of the stimulated subnetwork was associated with greater symptom improvement following DLPFC rTMS (p=0.013, figure 2, table S3), while multilayer BC of the equivalent DLPFC network in the vertex group did not predict treatment outcome (p=0.143, figure 2, table S3). Multilayer BC of the stimulated subnetwork in the preSMA group also did not predict treatment outcome (p=0.639, figure 2, table S3). No other associations between stimulation location or stimulated network graph features and treatment outcome survived correction for multiple comparisons (table S3). We found no significant associations between multilayer graph features of the stimulated region and treatment outcome in all participants combined.

**Figure 2:**
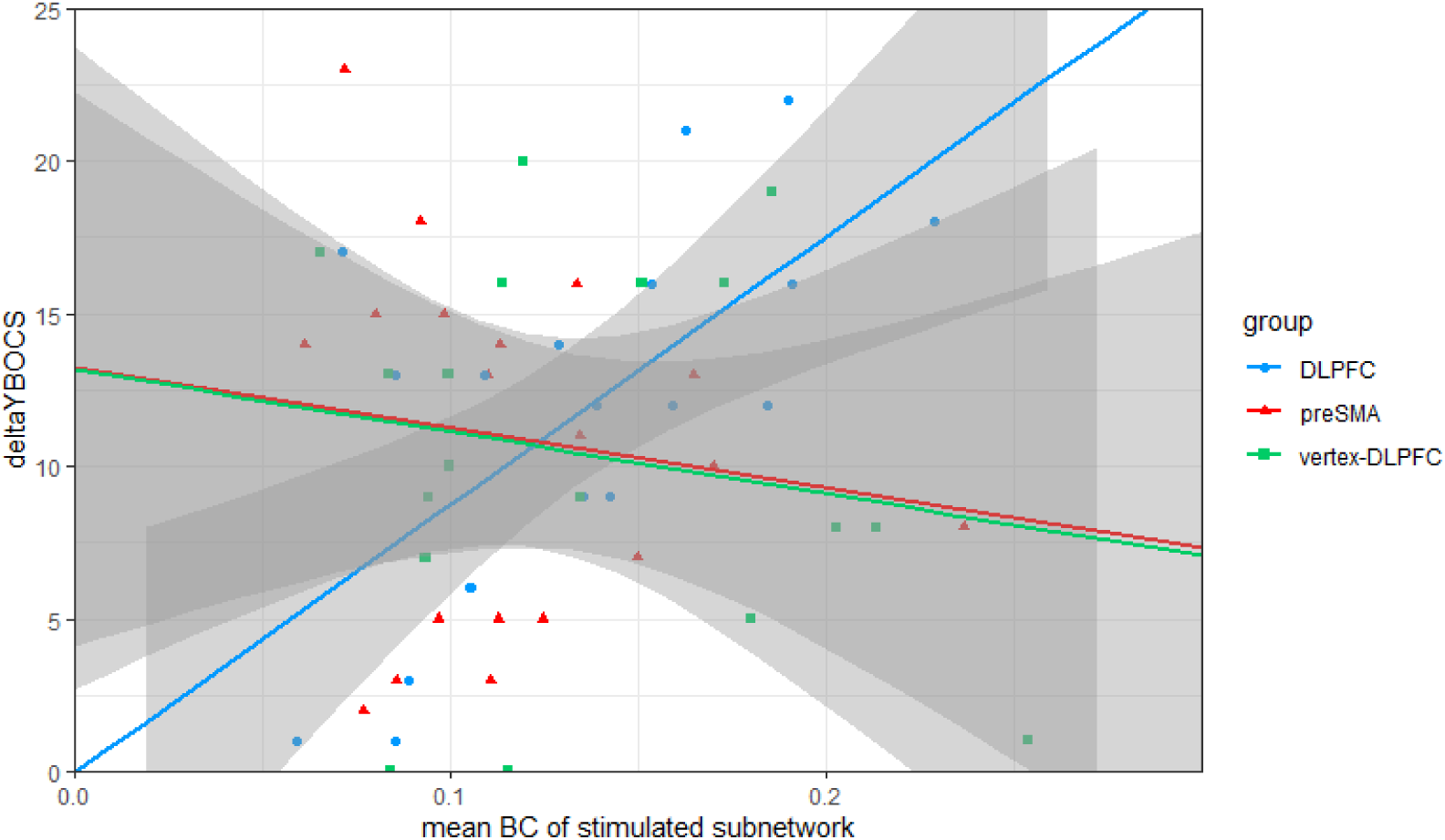
scatterplot showing association of mean BC of stimulated multilayer subnetwork (i.e. the Yeo subnetwork containing the DLPFC) with change in YBOCS scores in DLPFC group and vertex group.

In order to check whether association of BC with treatment outcome was specific to the stimulated network, we carried out further post hoc analyses within the DLPFC group. We calculated the association of treatment outcome with mean whole brain BC, mean whole brain BC without FPN and DMN, and mean sensorimotor network BC for the DLPFC group (Table S4). None were significantly associated with treatment outcome.

## Discussion

We investigated the relationship between single- and multilayer structural and functional brain network properties of the stimulation location and treatment outcome of a combined rTMS and ERP treatment for OCD. No associations were found between functional or structural single-layer graph metrics and treatment outcome for any group. For combined rTMS and ERP of the DLPFC, greater structural-functional multilayer BC of the stimulated subnetwork was associated with increased symptom improvement following treatment. This relationship was not observed in the equivalent DLPFC subnetwork in the vertex group, or in the stimulated subnetwork in the preSMA group.

Our findings indicate that structural-functional multilayer connectivity of the stimulated subnetwork may be a determinant of high frequency DLPFC rTMS effects in OCD: the more well integrated the stimulated subnetwork is with the rest of the brain, the greater the symptom improvement. The direction of effect is contrary to our hypothesised effect - we expected that lower global and higher local connectivity would be correlated with greater rTMS effect. Contrary to our other hypothesis, we did not observe an association with rTMS effect in the single-layer networks, while in our previous work on the functional single-layer brain network in healthy subjects[15] and OCD[16], we did find centrality measures that were predictive of rTMS outcome. One possible explanation for these observations is that these earlier studies were based on single-session *‘experimental’* rTMS, while the current study examines multi-session *‘therapeutic’* rTMS. There may be fundamental differences in working mechanisms between single and multi-session rTMS: a systematic review by Beynel et al[45] observed that single sessions of rTMS generally led to mixed increases and decreases in rsfMRI FC, while multiple sessions tended to lead to increases in FC, regardless of the stimulation frequency. This may be due to differences in plasticity mechanisms between single session and multisession rTMS, with changes following single sessions possibly reflecting short term plasticity, and following multisession rTMS compensatory/homeostatic plasticity[46–48].

Interestingly, we did not observe a relationship between mean multilayer betweenness centrality of the stimulated subnetwork and treatment outcome in the preSMA-stimulated group. This may be due to the fact that the most frequently stimulated resting state subnetwork differed between groups: in the DLPFC group this was the FPN, in the preSMA group the DMN. The FPN may be more relevant for rTMS treatment effect than the DMN: a study of DLPFC rTMS stimulation locations for the treatment for depression found that stimulation locations of responders had greater FC with the FPN, while the stimulation locations of nonresponders had greater FC with the DMN[49]. While our own analyses showed no difference in response rates between those stimulated at the FPN vs the DMN, our sample size is small and may be underpowered to detect this effect.

Our research gives an indication that neither structural nor functional connectivity alone, but rather the combined contribution of structural and functional networks is important for the efficacy of multi-session rTMS. Other research has also suggested that this interplay may be an important determinant of rTMS effects. TMS pulses have been shown to preferentially propagate via anatomical pathways of the structural network over those of the functional network[50], while at the same time a broad range of approaches and analyses indicate that the connectivity of the functional brain network is an important predictor of rTMS effect[6,7,9]. An integrative metric that combines these two brain features may allow a better representation of the complex relationship of both brain structure and function to brain stimulation outcome. For example, a study[51] of deep brain stimulation in depression found that a combined functional and structural connectivity mask predicted treatment outcome better than either functional or structural connectivity alone.

Our study has a number of weaknesses. As discussed in the primary outcomes paper for this trial[28], since we used low intensity active stimulation at the vertex, we cannot rule out the possibility that we induced some parietal stimulation during the treatment which may have had a therapeutic effect. The vertex condition is therefore an imperfect placebo; this finding requires replication with a true sham condition as a comparator. Our sample size per group is also relatively small: we may have failed to detect associations with the single-layer networks due to a lack of power (though the fact that we do detect an effect with multilayer networks with a small sample size and correction for multiple comparisons suggests that multilayer may be more sensitive to rTMS effects than single-layer). Our study also has several strengths. To our knowledge, this is the first study that uses multilayer networks to predict rTMS effect, and one of few that integrate both structural and functional neuroimaging data in the prediction of rTMS treatment outcomes. We use MST networks to construct our multilayer networks, removing potential bias related to link density or average connectivity[52].

Multilayer networks may offer a useful method to predict rTMS outcome and plan future treatments. Identifying properties of the stimulation location that are associated with greater treatment response may help improve selection of rTMS target sites. However, the true added value of this complex and labour-intensive approach needs to be evaluated before implementation in the clinic could be considered for response prediction and/or optimised targeting - a recent randomised trial[53] indicated that neuronavigation based on FC defined targets has limited added value over anatomical targeting.

In conclusion, we found that while structural and functional single-layer network properties were not associated with treatment outcome of rTMS for OCD, participants with greater multilayer BC of the stimulated subnetwork (greater integration of the stimulated subnetwork with the rest of the structural-functional network) showed greater improvement following DLPFC rTMS. Our results give a preliminary indication that the outcome of rTMS treatment for OCD is dependent on the combined contribution of structural and functional networks, and that multilayer network analysis may be a useful tool for future rTMS research.

## Data Availability

Data in the present study is not available to share

## Acknowledgements

This work was supported by a NWO-ZonMw VIDI grant (016.176.306) awarded to OAvdH, and a NWO VIDI grant (198.015) awarded to LD.

## Declaration of Competing Interest

The authors declare no competing interests.

## Abbreviations

rTMS: repetitive transcranial magnetic stimulation
OCD: obsessive-compulsive disorder
FC: functional connectivity
DLPFC: dorsolateral prefrontal cortex
dMRI: diffusion MRI
rsfMRI: resting state functional MRI
tbfMRI: task based functional MRI
ERP: exposure and response prevention
YBOCS: Yale Brown Obsessive Compulsive Scale
SRI: serotonin reuptake inhibitor
preSMA: pre-supplementary motor area
NS: node strength
BC: betweenness centrality
PC: participation coefficient
EC: eigenvector centrality
DC: degree centrality

## Notes

### Clinical Trial

NCT03667807

### Funding Statement

This study was funded by a NWO-ZonMw VIDI grant (016.176.306) awarded to OAvdH, and a NWO VIDI grant (198.015) awarded to LD.

### Author Declarations

Ethics committee/IRB of the Vrije Universiteit Medical Center, Amsterdam, gave ethical approval for this work

