## Supplement for "Structural-functional multilayer brain network properties of the stimulation location predict outcome of repetitive transcranial magnetic stimulation for obsessive-compulsive disorder"

**Supplementary information**

Supplementary Methods

1. *Interventions:*

   *rTMS/ERP intervention and blinding*

Randomization was stratified based on current use of SRI. Randomization was carried out following baseline assessments by a researcher not involved in clinical assessments. After randomization patients received either: 10Hz rTMS at 110% resting motor threshold (RMT) to the left DLPFC; 10Hz rTMS at 110% RMT to the left preSMA; or 10Hz rTMS at 60% RMT to the vertex. All participants received 3000 pulses in 30x10s trains with 30 seconds intertrain intervals[1].

The resting motor threshold (RMT) of the left motor cortex was determined once, within 2 weeks prior to starting rTMS treatment. Single TMS pulses were applied, gradually increasing in intensity, until five out of 10 pulses resulted in a right index finger motor-evoked potential of at least 50µV as measured using electromyography of the first dorsal interosseus muscle.

ERP sessions started within 10 minutes after the rTMS session was completed; each session lasted 60 minutes. Psychotherapists followed a protocolized treatment manual, including two introductory sessions, 16 exposure sessions (twice per week, directly following rTMS), one evaluation session, and one relapse prevention session, within ~10 weeks. Each patient worked with two different therapists during the treatment, and therapists received monthly supervision by an expert in CBT for OCD. Exposure sessions consisted of therapist-aided exposure exercises, and self-guided exposure exercises were assigned as between-session homework.

Participants, psychotherapists and clinical assessors were blinded to rTMS condition; rTMS therapists were unblinded but were instructed not to discuss the treatment conditions with participants or psychotherapists and did not take part in clinical assessments or ERP sessions.

*Creation of stimulation targets*

Please see Fitzsimmons et al[2] for full details of fMRI acquisition and processing for rTMS target creation, and for a full description of the Tower of London and Stop-Signal task cognitive tasks. For the DLPFC condition, the peak voxel in the left DLPFC (defined as Brodmann areas 9 and 46[3]) during the planning contrast of the Tower of London task (all planning conditions > baseline) was determined, and the stimulation target placed on the closest gyrus in the participant's T1 anatomical scan. For the preSMA condition, the local maximum of activation in the left preSMA (defined as the medial gyrus of Brodmann area 6 anterior to the anterior commissure[4] during the response inhibition contrast (successful stop trials > successful go trials) of the stop-signal task were used to determine the stimulation target for the preSMA condition. If no suitable local maximum could be located for the DLPFC or preSMA groups at a statistical threshold of P<0.01 uncorrected, the following literature coordinates (in MNI space) were used: i. Left DLPFC: -40, 28, 30[5]ii. Left preSMA: -4, 14, 58[6] For all participants in the vertex group the following MNI coordinates were used: 0, -34, 72[1] Once the individualized stimulation coordinate was defined, a 5mm ROI was created, warped from MNI to subject space and overlaid on a T1 MRI scan of the individual participant, allowing navigation to the individualized stimulation location at every rTMS treatment session using the Localite neuronavigation system (Localite GmbH, Bonn, Germany)

### *2. Detailed fMRIprep preprocessing boilerplate for rsfMRI data*

Results included in this manuscript come from preprocessing performed using fMRIPrep 21.0.1[7,8]; RRID:SCR_016216, which is based on Nipype 1.6.1[9,10]; RRID:SCR_002502.

*Preprocessing of B0 inhomogeneity mappings*

A total of 2 fieldmaps were available per subject. A B0-nonuniformity map (or fieldmap) was estimated based on two (or more) echo-planar imaging (EPI) references with topup ([11]; FSL 6.0.5.1:57b01774).

*Anatomical data preprocessing*

A total of 2 T1-weighted (T1w) images were found within the input BIDS dataset. All of them were corrected for intensity non-uniformity (INU) with N4BiasFieldCorrection [12], distributed with ANTs 2.3.3 ([13], RRID:SCR_004757). The T1w-reference was then skull-stripped with a Nipype implementation of the antsBrainExtraction.sh workflow (from ANTs), using OASIS30ANTs as target template. Brain tissue segmentation of cerebrospinal fluid (CSF), white-matter (WM) and gray-matter (GM) was performed on the brain-extracted T1w using fast (FSL 6.0.5.1:57b01774, RRID:SCR_002823, [14]). A T1w-reference map was computed after registration of 2 T1w images (after INU-correction) using mri_robust_template (FreeSurfer 6.0.1, [15]). Brain surfaces were reconstructed using recon-all (FreeSurfer 6.0.1, RRID:SCR_001847,[16]), and the brain mask estimated previously was refined with a custom variation of the method to reconcile ANTs-derived and FreeSurfer-derived segmentations of the cortical gray-matter of Mindboggle (RRID:SCR_002438,[17]). Volume-based spatial normalization to two standard spaces (MNI152NLin6Asym, MNI152NLin2009cAsym) was performed through nonlinear registration with antsRegistration (ANTs 2.3.3), using brain-extracted versions of both T1w reference and the T1w template. The following templates were selected for spatial normalization: FSL's MNI ICBM 152 non-linear 6th Generation Asymmetric Average Brain Stereotaxic Registration Model [[18], RRID:SCR_002823; TemplateFlow ID: MNI152NLin6Asym], ICBM 152 Nonlinear Asymmetrical template version 2009c [[19], RRID:SCR_008796; TemplateFlow ID: MNI152NLin2009cAsym].

*Functional data preprocessing*

For each of the 4 BOLD runs per subject (across all tasks and sessions), the following preprocessing was performed. First, a reference volume and its skull-stripped version were generated using a custom methodology of fMRIPrep. Head-motion parameters with respect to the BOLD reference (transformation matrices, and six corresponding rotation and translation parameters) are estimated before any spatiotemporal filtering using mcflirt (FSL 6.0.5.1:57b01774,[20]). The estimated fieldmap was then aligned with rigid-registration to the target EPI (echo-planar imaging) reference run. The field coefficients were mapped on to the reference EPI using the transform. BOLD runs were slice-time corrected to 1.07s (0.5 of slice acquisition range 0s-2.15s) using 3dTshift from AFNI ([21], RRID:SCR_005927). The BOLD reference was then co-registered to the T1w reference using bbregister (FreeSurfer) which implements boundary-based registration[22]. Co-registration was configured with six degrees of freedom. Several confounding time-series were calculated based on the preprocessed BOLD: framewise displacement (FD), DVARS and three region-wise global signals. FD was computed using two formulations following Power (absolute sum of relative motions,[23]) and Jenkinson (relative root mean square displacement between affines,[20]). FD and DVARS are calculated for each functional run, both using their implementations in Nipype (following the definitions by [23]). The three global signals are extracted within the CSF, the WM, and the whole-brain masks. Additionally, a set of physiological regressors were extracted to allow for component-based noise correction (CompCor,[24]). Principal components are estimated after high-pass filtering the preprocessed BOLD time-series (using a discrete cosine filter with 128s cut-off) for the two CompCor variants: temporal (tCompCor) and anatomical (aCompCor). tCompCor components are then calculated from the top 2% variable voxels within the brain mask. For aCompCor, three probabilistic masks (CSF, WM and combined CSF+WM) are generated in anatomical space. The implementation differs from that of Behzadi et al. in that instead of eroding the masks by 2 pixels on BOLD space, the aCompCor masks are subtracted a mask of pixels that likely contain a volume fraction of GM. This mask is obtained by dilating a GM mask extracted from the FreeSurfer’s aseg segmentation, and it ensures components are not extracted from voxels containing a minimal fraction of GM. Finally, these masks are resampled into BOLD space and binarized by thresholding at 0.99 (as in the original implementation). Components are also calculated separately within the WM and CSF masks. For each CompCor decomposition, the k components with the largest singular values are retained, such that the retained components’ time series are sufficient to explain 50 percent of variance across the nuisance mask (CSF, WM, combined, or temporal). The remaining components are dropped from consideration. The head-motion estimates calculated in the correction step were also placed within the corresponding confounds file. The confound time series derived from head motion estimates and global signals were expanded with the inclusion of temporal derivatives and quadratic terms for each [25]. Frames that exceeded a threshold of 0.5 mm FD or 1.5 standardised DVARS were annotated as motion outliers. The BOLD time-series were resampled into standard space, generating a preprocessed BOLD run in MNI152NLin6Asym space. First, a reference volume and its skull-stripped version were generated using a custom methodology of fMRIPrep. The BOLD time-series were resampled onto the following surfaces (FreeSurfer reconstruction nomenclature): fsnative, fsaverage5. Automatic removal of motion artifacts using independent component analysis (ICA-AROMA, [26]) was performed on the preprocessed BOLD on MNI space time-series after removal of non-steady state volumes and spatial smoothing with an isotropic, Gaussian kernel of 6mm FWHM (full-width half-maximum). Corresponding “non-aggresively” denoised runs were produced after such smoothing. Additionally, the “aggressive” noise-regressors were collected and placed in the corresponding confounds file. All resamplings can be performed with a single interpolation step by composing all the pertinent transformations (i.e. head-motion transform matrices, susceptibility distortion correction when available, and co-registrations to anatomical and output spaces). Gridded (volumetric) resamplings were performed using antsApplyTransforms (ANTs), configured with Lanczos interpolation to minimize the smoothing effects of other kernels [27]. Non-gridded (surface) resamplings were performed using mri_vol2surf (FreeSurfer).

Many internal operations of fMRIPrep use Nilearn 0.8.1 ([28], RRID:SCR_001362), mostly within the functional processing workflow. For more details of the pipeline, see the section corresponding to workflows in fMRIPrep’s documentation.

Copyright Waiver

The above boilerplate text was automatically generated by fMRIPrep with the express intention that users should copy and paste this text into their manuscripts unchanged. It is released under the CC0 license.

*3. Excluded Schaefer 300 atlas[29] regions*

7Networks_LH_Limbic_OFC_3

7Networks_LH_Limbic_TempPole_1

7Networks_LH_Limbic_TempPole_4

7Networks_LH_Cont_OFC_1

7Networks_RH_Limbic_OFC_3

7Networks_RH_Limbic_TempPole_1

7Networks_RH_Limbic_TempPole_4

[13] Avants B, Epstein C, Grossman M, Gee J. Symmetric diffeomorphic image registration with

cross-correlation: Evaluating automated labeling of elderly and neurodegenerative brain. Medical Image Analysis 2008;12:26–41. https://doi.org/10.1016/j.media.2007.06.004.
[14] Zhang Y, Brady M, Smith S. Segmentation of brain MR images through a hidden Markov random field model and the expectation-maximization algorithm. IEEE Trans Med Imaging 2001;20:45–57. https://doi.org/10.1109/42.906424.
[15] Reuter M, Rosas HD, Fischl B. Highly accurate inverse consistent registration: A robust approach. NeuroImage 2010;53:1181–96. https://doi.org/10.1016/j.neuroimage.2010.07.020.
[16] Dale AM, Fischl B, Sereno MI. Cortical Surface-Based Analysis. NeuroImage 1999;9:179–94. https://doi.org/10.1006/nimg.1998.0395.
[17] Klein A, Ghosh SS, Bao FS, Giard J, Häme Y, Stavsky E, et al. Mindboggling morphometry of human brains. PLoS Comput Biol 2017;13:e1005350. https://doi.org/10.1371/journal.pcbi.1005350.
[18] Evans AC, Janke AL, Collins DL, Baillet S. Brain templates and atlases. NeuroImage 2012;62:911–22. https://doi.org/10.1016/j.neuroimage.2012.01.024.
[19] Fonov V, Evans A, McKinstry R, Almli C, Collins D. Unbiased nonlinear average age-appropriate brain templates from birth to adulthood. NeuroImage 2009;47:S102. https://doi.org/10.1016/S1053-8119(09)70884-5.
[20] Jenkinson M, Bannister P, Brady M, Smith S. Improved optimization for the robust and accurate linear registration and motion correction of brain images. Neuroimage 2002;17:825–41.
[21] Cox RW, Hyde JS. Software tools for analysis and visualization of fMRI data. NMR Biomed 1997;10:171–8. https://doi.org/10.1002/(SICI)1099-1492(199706/08)10:4/5<171::AID-NBM453>3.0.CO;2-L.
[22] Greve DN, Fischl B. Accurate and robust brain image alignment using boundary-based registration. NeuroImage 2009;48:63–72. https://doi.org/10.1016/j.neuroimage.2009.06.060.
[23] Power JD, Mitra A, Laumann TO, Snyder AZ, Schlaggar BL, Petersen SE. Methods to detect, characterize, and remove motion artifact in resting state fMRI. NeuroImage 2014;84:320–41. https://doi.org/10.1016/j.neuroimage.2013.08.048.
[24] Behzadi Y, Restom K, Liau J, Liu TT. A component based noise correction method (CompCor) for BOLD and perfusion based fMRI. NeuroImage 2007;37:90–101. https://doi.org/10.1016/j.neuroimage.2007.04.042.
[25] Satterthwaite TD, Elliott MA, Gerraty RT, Ruparel K, Loughead J, Calkins ME, et al. An improved framework for confound regression and filtering for control of motion artifact in the preprocessing of resting-state functional connectivity data. NeuroImage 2013;64:240–56. https://doi.org/10.1016/j.neuroimage.2012.08.052.
[26] Pruim RHR, Mennes M, Van Rooij D, Llera A, Buitelaar JK, Beckmann CF. ICA-AROMA: A robust ICA-based strategy for removing motion artifacts from fMRI data. NeuroImage 2015;112:267–77. https://doi.org/10.1016/j.neuroimage.2015.02.064.
[27] Lanczos C. Evaluation of Noisy Data. Journal of the Society for Industrial and Applied Mathematics Series B Numerical Analysis 1964;1:76–85. https://doi.org/10.1137/0701007.
[28] Abraham A, Pedregosa F, Eickenberg M, Gervais P, Mueller A, Kossaifi J, et al. Machine learning for neuroimaging with scikit-learn. Front Neuroinform 2014;8. https://doi.org/10.3389/fninf.2014.00014.
[29] Schaefer A, Kong R, Gordon EM, Laumann TO, Zuo X-N, Holmes AJ, et al. Local-Global Parcellation of the Human Cerebral Cortex from Intrinsic Functional Connectivity MRI. Cereb Cortex 2018;28:3095–114. https://doi.org/10.1093/cercor/bhx179.

Supplementary tables

Table S1: results of multiple regressions predicting clinical outcome from graph properties of stimulation locations derived from single-layer structural networks. Coefficients for T0-T2 delta YBOCS, adjusted for age, sex and T0 YBOCS score, are shown.

| group | Predictor | Coefficient | Standard Error | P value | Adjusted R2 for model | P value of model | group | Predictor | Coefficient | Standard Error | P value | Adjusted R2 for model | P value of model |
|---|---|---|---|---|---|---|---|---|---|---|---|---|---|
| DLPFC | BC of stimulated node | 0.001 | 0.001 | 0.255 | -0.064 | 0.586 | Vertex-DLPFC | BC of stimulated node | 0.001 | 0.001 | 0.441 | 0.033 | 0.378 |
| | PC of stimulated node | 0.007 | 0.005 | 0.206 | -0.097 | 0.667 | | PC of stimulated node | -0.004 | 0.008 | 0.589 | 0.129 | 0.226 |
| | ECof stimulated node | 0.000 | 0.001 | 0.882 | -0.261 | 0.990 | | ECof stimulated node | 0.001 | 0.001 | 0.275 | 0.348 | 0.046 |
| | NS of stimulated node | 0.000 | 0.000 | 0.365 | -0.052 | 0.557 | | NS of stimulated node | 0.000 | 0.000 | 0.375 | 0.047 | 0.354 |
| | Mean BC of stimulated network | 0.000 | 0.000 | 0.876 | -0.062 | 0.581 | | Mean BC of stimulated network | 0.000 | 0.000 | 0.168 | -0.091 | 0.639 |
| | Mean PC of stimulated network | 0.000 | 0.000 | 0.225 | 0.103 | 0.251 | | Mean PC of stimulated network | 0.000 | 0.000 | 0.076 | 0.000 | 0.442 |
| | Mean EC of stimulated network | 0.000 | 0.002 | 0.900 | -0.131 | 0.751 | | Mean EC of stimulated network | 0.004 | 0.002 | 0.056 | 0.220 | 0.126 |
| | Mean NS of stimulated network | 0.000 | 0.000 | 0.470 | -0.068 | 0.595 | | **Mean NS of stimulated network** | **0.000** | **0.000** | **0.036*** | **0.093** | **0.278** |
| preSMA | BC of stimulated node | 0.000 | 0.001 | 0.861 | 0.197 | 0.112 | Vertex-preSMA | BC of stimulated node | -0.001 | 0.002 | 0.535 | -0.051 | 0.549 |
| | PC of stimulated node | -0.003 | 0.003 | 0.322 | -0.103 | 0.713 | | PC of stimulated node | 0.002 | 0.004 | 0.564 | 0.139 | 0.213 |
| | ECof stimulated node | 0.000 | 0.001 | 0.896 | -0.025 | 0.499 | | ECof stimulated node | 0.000 | 0.001 | 0.720 | 0.161 | 0.186 |
| | NS of stimulated node | 0.000 | 0.000 | 0.850 | -0.104 | 0.717 | | NS of stimulated node | -0.001 | 0.000 | 0.124 | 0.223 | 0.124 |
| | Mean BC of stimulated network | 0.000 | 0.000 | 0.664 | -0.035 | 0.526 | | Mean BC of stimulated network | 0.000 | 0.000 | 0.471 | 0.006 | 0.431 |
| | Mean PC of stimulated network | 0.000 | 0.000 | 0.821 | -0.102 | 0.710 | | Mean PC of stimulated network | 0.000 | 0.000 | 0.338 | -0.084 | 0.625 |
| | Mean EC of stimulated network | -0.001 | 0.002 | 0.732 | -0.038 | 0.532 | | Mean EC of stimulated network | 0.001 | 0.002 | 0.607 | -0.148 | 0.770 |
| | Mean NS of stimulated network | 0.000 | 0.000 | 0.791 | -0.234 | 0.994 | | **Mean NS of stimulated network** | **0.000** | **0.000** | **0.049*** | **0.194** | **0.150** |

BC, betweenness centrality; PC, participation coefficient; EC, eigenvector centrality; NS, node strength; DLPFC, dorsolateral prefrontal cortex; preSMA, pre-supplementary motor area; *p<0.05 but does not survive correction for multiple comparisons with α=0.019

Table S2: results of multiple regressions predicting clinical outcome from graph properties of stimulation locations derived from single-layer functional networks. Coefficients for T0-T2 delta YBOCS, adjusted for age, sex and T0 YBOCS score, are shown.

| group | Predictor | Coefficient | Standard Error | P value | Adjusted R2 for model | P value of model | group | Predictor | Coefficient | Standard Error | P value | Adjusted R2 for model | P value of model |
|---|---|---|---|---|---|---|---|---|---|---|---|---|---|
| DLPFC | BC of stimulated node | 0.000 | 0.000 | 0.458 | 0.210 | 0.123 | Vertex-DLPFC | BC of stimulated node | 0.000 | 0.000 | 0.160 | -0.001 | 0.444 |
| | PC of stimulated node | -0.001 | 0.002 | 0.736 | -0.107 | 0.692 | | PC of stimulated node | 0.004 | 0.003 | 0.256 | -0.066 | 0.583 |
| | ECof stimulated node | 0.000 | 0.000 | 0.477 | 0.062 | 0.318 | | EC of stimulated node | 0.001 | 0.001 | 0.086 | 0.260 | 0.095 |
| | NS of stimulated node | 0.001 | 0.002 | 0.498 | -0.180 | 0.864 | | NS of stimulated node | 0.003 | 0.002 | 0.155 | 0.158 | 0.189 |
| | Mean BC of stimulated network | 0.000 | 0.000 | 0.634 | 0.085 | 0.279 | | Mean BC of stimulated network | 0.000 | 0.000 | 0.500 | 0.014 | 0.415 |
| | Mean PC of stimulated network | 0.000 | 0.000 | 0.525 | 0.058 | 0.325 | | Mean PC of stimulated network | 0.000 | 0.000 | 0.265 | 0.000 | 0.443 |
| | Mean EC of stimulated network | -0.001 | 0.002 | 0.559 | -0.206 | 0.916 | | Mean EC of stimulated network | 0.002 | 0.001 | 0.152 | 0.165 | 0.181 |
| | Mean NS of stimulated network | 0.001 | 0.001 | 0.421 | -0.206 | 0.916 | | Mean NS of stimulated network | 0.000 | 0.001 | 0.792 | -0.209 | 0.895 |
| preSMA | BC of stimulated node | 0.000 | 0.000 | 0.417 | -0.056 | 0.582 | Vertex-preSMA | BC of stimulated node | 0.000 | 0.000 | 0.821 | 0.066 | 0.320 |
| | PC of stimulated node | -0.002 | 0.002 | 0.515 | -0.145 | 0.830 | | PC of stimulated node | 0.003 | 0.003 | 0.239 | -0.126 | 0.720 |
| | EC of stimulated node | 0.000 | 0.000 | 0.720 | -0.035 | 0.525 | | ECof stimulated node | 0.001 | 0.001 | 0.271 | -0.064 | 0.579 |
| | NS of stimulated node | 0.001 | 0.002 | 0.362 | 0.066 | 0.294 | | NS of stimulated node | 0.002 | 0.003 | 0.547 | -0.043 | 0.531 |
| | Mean BC of stimulated network | 0.000 | 0.000 | 0.601 | -0.103 | 0.714 | | Mean BC of stimulated network | 0.000 | 0.000 | 0.083 | 0.271 | 0.087 |
| | Mean PC of stimulated network | 0.000 | 0.000 | 0.333 | -0.080 | 0.649 | | Mean PC of stimulated network | 0.000 | 0.000 | 0.336 | -0.037 | 0.519 |
| | Mean EC of stimulated network | -0.001 | 0.001 | 0.525 | -0.101 | 0.707 | | Mean EC of stimulated network | 0.001 | 0.002 | 0.637 | 0.015 | 0.414 |
| | Mean NS of stimulated network | 0.001 | 0.001 | 0.554 | -0.182 | 0.918 | | Mean NS of stimulated network | 0.001 | 0.003 | 0.846 | -0.150 | 0.773 |

BC, betweenness centrality; PC, participation coefficient; EC, eigenvector centrality; NS, node strength; DLPFC, dorsolateral prefrontal cortex; preSMA, pre-supplementary motor area

Table S3: results of multiple regressions predicting clinical outcome from graph properties of stimulation locations derived from multilayer functional-structural networks. Coefficients for T0-T2 delta YBOCS, adjusted for age, sex and T0 YBOCS score, are shown.

| group | Predictor | Coefficient | Standard Error | P value | Adjusted R2 for model | P value of model | group | Predictor | Coefficient | Standard Error | P value | Adjusted R2 for model | P value of model |
|---|---|---|---|---|---|---|---|---|---|---|---|---|---|
| DLPFC | BC of stimulated node | 0.005 | 0.004 | 0.199 | -0.108 | 0.693 | Vertex-DLPFC | BC of stimulated node | 0.003 | 0.008 | 0.708 | -0.134 | 0.738 |
| | EC of stimulated node | 0.004 | 0.005 | 0.417 | -0.131 | 0.752 | | EC of stimulated node | -0.001 | 0.002 | 0.583 | -0.158 | 0.791 |
| | DC of stimulated node | 0.005 | 0.004 | 0.175 | -0.004 | 0.449 | | DC of stimulated node | 0.003 | 0.008 | 0.734 | -0.062 | 0.574 |
| | **Mean BC of stimulated network** | **0.004** | **0.002** | **0.013**** | **0.391** | **0.025** | | Mean BC of stimulated network | -0.004 | 0.002 | 0.143 | 0.176 | 0.169 |
| | Mean EC of stimulated network | 0.002 | 0.002 | 0.225 | -0.116 | 0.714 | | Mean EC of stimulated network | -0.004 | 0.002 | 0.053 | 0.109 | 0.253 |
| | Mean DC of stimulated network | 0.002 | 0.002 | 0.325 | 0.035 | 0.368 | | Mean DC of stimulated network | -0.003 | 0.003 | 0.291 | 0.046 | 0.356 |
| preSMA | BC of stimulated node | -0.004 | 0.004 | 0.335 | -0.122 | 0.767 | Vertex-preSMA | BC of stimulated node | 0.000 | 0.006 | 0.948 | -0.140 | 0.752 |
| | EC of stimulated node | -0.003 | 0.004 | 0.500 | 0.002 | 0.432 | | EC of stimulated node | 0.002 | 0.005 | 0.741 | 0.013 | 0.416 |
| | DC of stimulated node | 0.003 | 0.003 | 0.364 | -0.053 | 0.574 | | DC of stimulated node | 0.002 | 0.005 | 0.679 | 0.311 | 0.063 |
| | Mean BC of stimulated network | -0.001 | 0.001 | 0.639 | -0.134 | 0.800 | | Mean BC of stimulated network | -0.001 | 0.002 | 0.562 | 0.068 | 0.318 |
| | Mean EC of stimulated network | 0.002 | 0.002 | 0.290 | -0.158 | 0.861 | | Mean EC of stimulated network | -0.003 | 0.002 | 0.117 | 0.026 | 0.392 |
| | Mean DC of stimulated network | 0.002 | 0.003 | 0.445 | -0.066 | 0.609 | | Mean DC of stimulated network | -0.001 | 0.003 | 0.597 | 0.001 | 0.441 |

BC, betweenness centrality; EC, eigenvector centrality; DC, degree centrality; DLPFC, dorsolateral prefrontal cortex; preSMA, pre-supplementary motor area; **=survives correction for multiple comparisons with p<0.025

Table S4: results of post hoc multiple regressions predicting clinical outcome from graph properties of stimulation locations derived from multilayer functional-structural networks in the DLPFC group. Coefficients for T0-T2 delta YBOCS, adjusted for age, sex and T0 YBOCS score, are shown.

| group | Predictor | Coefficient | Standard Error | P value | Adjusted R2 for model | P value of model |
|---|---|---|---|---|---|---|
| DLPFC | Wholebrain mean BC | 0.003 | 0.001 | 0.076 | 0.196 | 0.136 |
| | Wholebrain mean BC without DMN and FPN | 0.002 | 0.001 | 0.175 | 0.101 | 0.254 |
| | Mean BC of somatomotor network | 0.000 | 0.001 | 0.747 | 0.002 | 0.436 |

BC, betweenness centrality; DMN, default mode network; FPN, frontoparietal network
